# Benchmarking methods integrating GWAS and single-cell transcriptomic data for mapping trait-cell type associations

**DOI:** 10.1101/2025.05.24.25328275

**Authors:** Ang Li, Peter Allen, Yong Wang, Haochen Cui, Tian Lin, Yuliangzi Sun, Xuemin Wang, Xiao Tan, Alicia Walker, Siqi Wang, Zheng Yao, Ruolan Zhao, Jian Yang, Shuyang Yao, Patrick F. Sullivan, Jens Hjerling-Leffler, Naomi R. Wray, Jian Zeng

## Abstract

Genome-wide association studies (GWAS) have discovered numerous trait-associated variants, but their biological context remains unclear. Integrating GWAS summary statistics with single-cell RNA-sequencing (scRNA-seq) data enables prioritization of cell types in which these variants influence traits. Existing methods broadly follow two strategies: “single cell to GWAS”, which identifies cell-type-specific genes and tests their enrichment in GWAS signals, and “GWAS to single cell”, which begins with GWAS-prioritized genes and scores cells or cell types according to their expression profiles. Here, we developed a literature-informed benchmark by integrating PubMed evidence with large language model-assisted literature synthesis to evaluate 20 trait-cell type mapping methods. We identify CATCH, a Cauchy combination of complementary methods, as the most robust overall approach, consistently achieving high statistical power while maintaining effective false-positive control across simulations and real-data benchmarks. We further identify key determinants of performance, including cell-type specificity metrics, GWAS statistical power, and the diversity of scRNA-seq reference datasets, providing practical guidance for the development and application of trait-cell type mapping methods.

## Introduction

Genetic variants likely affect complex traits and diseases by acting at the level of specific cell types^1–5^. Identifying these cell types is essential for understanding disease aetiology and for developing new preventative and therapeutic approaches for personalized medicine^6–8^. A powerful approach is to integrate two complementary data modalities: summary statistics from genome-wide association studies (GWAS), which capture SNP-trait associations, and gene transcript-by-cell matrices from single-cell or single-nucleus RNA sequencing (both abbreviated as scRNA-seq hereafter), from which cell-type-specific gene expression profiles can be derived. In principle, a cell type is prioritized if GWAS signals are significantly associated with the gene expression specificity of the cell type. Methods that integrate GWAS and scRNA-seq data to prioritize trait-associated cell types, referred to as trait-cell type mapping methods, have proliferated rapidly. As of June 2026, we identified more than 16 computational methods^9–19^ for this purpose (**Supplementary Table 1**). However, their relative performance and the factors influencing it remain unclear.

Benchmarking these methods is challenging for several reasons. First, the true relationships between complex traits and cell types remain largely unknown, making it difficult to compare methods using real data. Currently, no gold standard exists for benchmarking, and new methods are often evaluated primarily by the number of trait-cell type associations they identify relative to previous approaches, a strategy that may favour methods with inflated false discovery rates^17,20^. Second, while GWAS datasets have grown rapidly in sample size over the past decade, statistical power varies widely across traits^21,22^, which may lead to different methods performing optimally for different traits. Third, scRNA-seq datasets are increasingly available across different species, conditions (e.g., cases versus controls), longitudinal time points, organs, and tissue regions, presenting opportunities for discovery but also challenges in effectively integrating and analysing multiple datasets^23^. Methods differ in how they identify gene sets associated with relative cell type specificity^24,25^, and these gene sets can vary depending on experimental conditions, sequencing technology/depth, biological contexts, and data sources^26^.

Existing trait-cell type mapping methods can be classified into two strategies based on their conceptual distinctions (**Fig. 1**). From single cell transcriptomics to GWAS (SC-to-GWAS) methods first identify specifically expressed genes (SEGs) for each cell type and then test whether these genes are enriched for GWAS signals. Multiple methods have been proposed to identify SEGs^13,16,27–30^ (**Supplementary Table 2**), and the resulting annotations can subsequently be integrated with GWAS summary statistics using approaches such as stratified LD score regression^10,11^ (sLDSC) or MAGMA gene-set enrichment analysis ^20,31,32^ (MAGMA-GSEA) and gene-property analysis^12,32^(MAGMA-GPA). In contrast, from GWAS to single cell transcriptomics (GWAS-to-SC) methods start with GWAS-prioritized genes and compute a disease relevance score for each individual cell — the weighted aggregate expression of these genes, with scDRS^9^ being a representative method.

**Figure 1.**
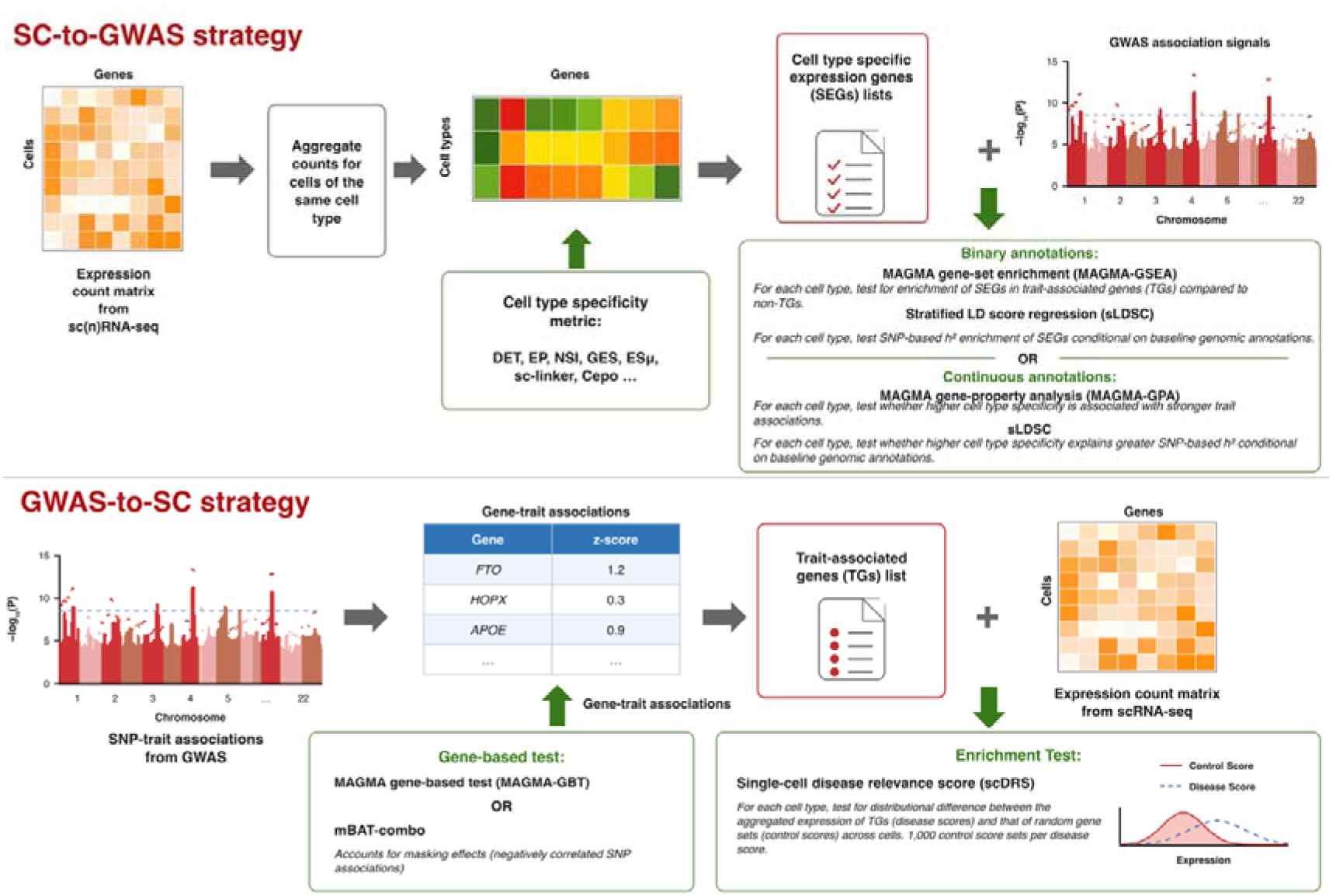
Two main strategies for integrating GWAS with single cell transcriptomic data to prioritize trait-associated cell types. The “single cell (SC)-to-GWAS” strategy (representing most methods) identifies gene sets with cell-type-specific expression and then follows with enrichment analyses applied to GWAS summary statistics. Conversely, the “GWAS-to-SC” strategy begins with a list of trait-associated genes and calculates a cumulative disease score per cell based on gene expression count data. Descriptions of the methods in **Supplementary Tables 1** and **2**.

To address the lack of established benchmarks for trait-cell type mapping, we developed a literature-informed benchmarking framework that combines PubMed evidence with assessments from 12 large language models (LLMs), thereby leveraging existing biomedical knowledge while reducing publication bias. Recent advances have demonstrated that LLMs can reliably synthesize biomedical knowledge across a range of clinical and computational biology applications^33–36^. Here, we leverage LLM-assisted literature synthesis to construct a benchmark for trait-cell type mapping while systematically evaluating the utility and robustness of LLMs for this application.

In this study, we present a comprehensive benchmarking framework comprising controlled simulations and an LLM-assisted literature-informed benchmark based on real data. We systematically evaluate 20 methods across SC-to-GWAS and GWAS-to-SC strategies (**Supplementary Table 3**), and propose CATCH, an evidence-based Cauchy combination framework^37,38^ that integrates complementary methods across these two strategies. We further investigate how factors such as cell-type specificity metrics, GWAS statistical power, and scRNA-seq reference selection influence method performance. Together, these analyses establish a robust benchmarking framework and provide practical guidance for integrating genetic and single-cell data to prioritize cell types for complex traits.

## Results

### Simulation-based benchmarking identifies a robust Cauchy combination strategy

We performed simulations using HapMap3-matched genotypes from 20,000 unrelated UK Biobank participants of inferred European ancestry, in which pancreatic β-cells were specified as the causal cell type (**Fig. 2a; Methods**). Simulations were based on a human atlas scRNA-seq dataset ^39^ comprising 155 cell types across multiple organs and a pancreatic endocrine-islet scRNA-seq dataset^40^ comprising ten cell types. To avoid favouring any particular cell-type specificity metric evaluated in this study, β-cell-specific genes were defined using an externally curated list of 94 established β-cell marker genes^41,42^(**Supplementary Table 4**). Four causal scenarios were simulated by assigning either 20% or 50% of causal variants to β-cell marker genes, each at a heritability (*h²*) of 0.1 or 0.2. We additionally simulated seven null scenarios without β-cell-specific genetic enrichment, including a no-effect null (*h²* = 0), randomly distributed causal variants, functional-annotation-enriched causal variants, and infinitesimal genetic architectures, each at *h²* = 0.1 and 0.2. Each simulation scenario was repeated 30 times. The simulations provided a controlled setting for evaluating statistical power and family-wise error rate (FWER) under the null.

**Figure 2.**
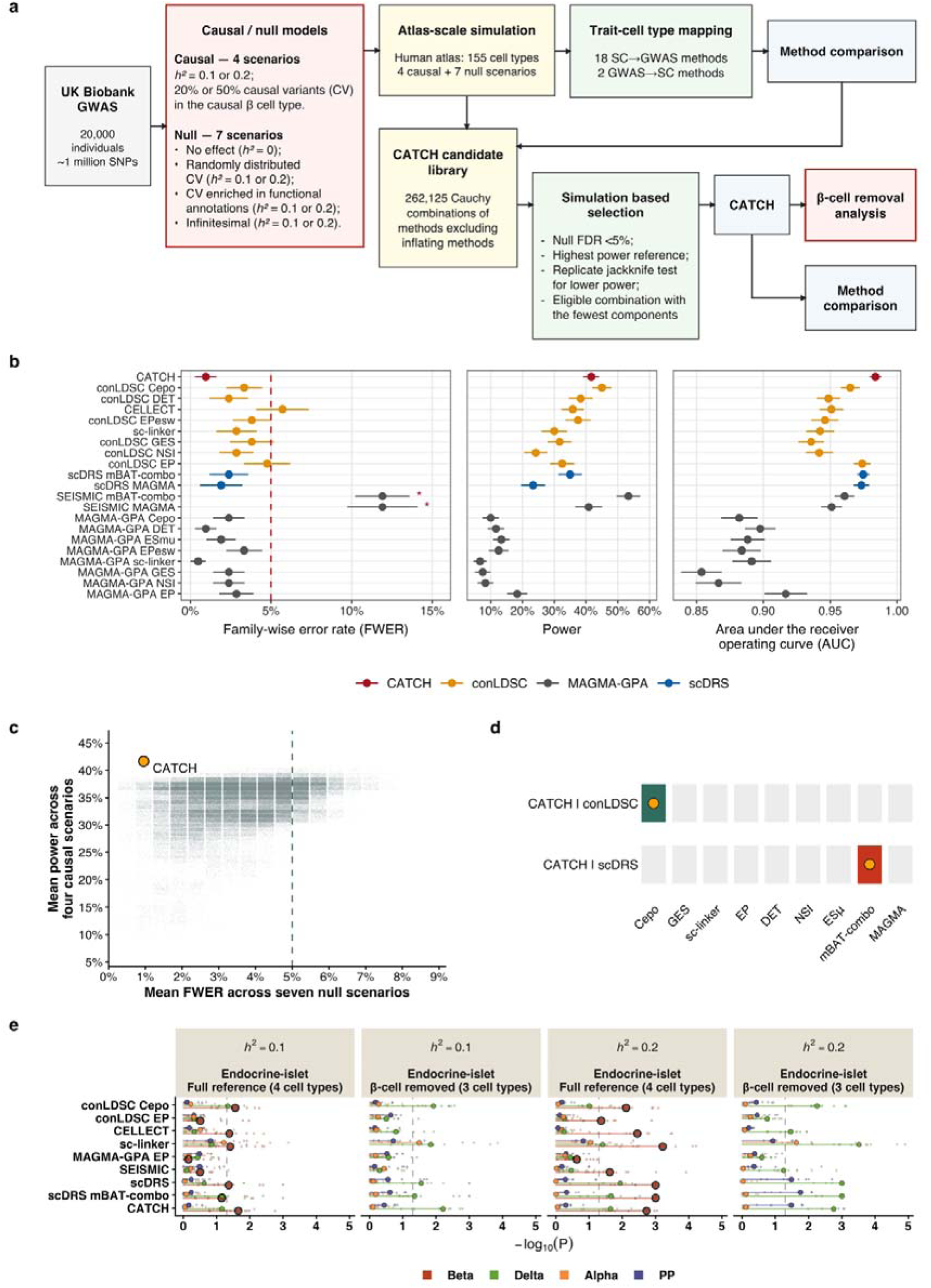
Simulation-based benchmarking of 20 method configurations identifies a CATCH strategy for trait-cell type mapping. **(a)** Overview of the simulation framework using real genotype data and human atlas scRNA-seq data to benchmark method and identify CATCH. **(b)** Performance of 20 method configurations and CATCH across four causal and seven null simulation scenarios (30 replicates per scenario) using a human atlas reference of 155 cell types. Family-wise error rate (FWER) is the proportion of null simulation replicates with at least one Benjamini-Hochberg (BH)-significant cell type (BH-adjusted P<0.05), averaged across the seven null scenarios. Power is the proportion of causal simulation replicates in which the causal pancreatic β cell is detected, averaged across the four causal scenarios. For calculation of s.e.m. and statistical tests, performance was first averaged across the relevant scenarios within each replicate. Points and horizontal bars show the mean□±□s.e.m. across 30 replicates. Dashed red line indicates FWER□=□0.05. Red asterisks indicate P<0.05 from one-sided t-tests comparing each method’s replicate-level FWER with 0.05. Colours denote CATCH (red), conLDSC-based configurations (orange), MAGMA-GPA configurations (grey), and scDRS configurations (blue). All methods used symmetric ±100-kb SNP-to-gene windows. **(c)** Power and FWER for 262,125 candidate CATCH configurations evaluated across the same simulation scenarios. Each point represents one configuration. The dashed line indicates FWER=0.05. CATCH was selected as the smallest well-calibrated combination whose power was not significantly lower than the maximum observed power (**Methods**). **(d)** Components of the final CATCH configuration: conLDSC-Cepo and scDRS-mBAT-combo. **(e**) Performance after removing the causal β cells from a four-cell-type endocrine-islet reference. Association signals before and after β-cell removal are shown as -log10(P). Bars indicate the median top-cell signal, points indicate replicate-level values. Colours indicate the most frequently top-ranked cell type after β-cell removal.

We first compared all 20 benchmarked methods spanning the major methodological families (**Supplementary Table 3)**. These included representative published methods such as conLDSC-EP (sLDSC with expression proportion^27^ (EP) as a continuous annotation), MAGMA-GPA-EP^12^, CELLECT^13^, sc-linker^9^, scDRS^9^, and seismic^43^, together with additional combinations generated using alternative cell-type specificity metrics (e.g., DET^44^, ES_µ_^13^, and Cepo^28^) and gene-based association methods (e.g., mBAT-combo^45^). Although these methods have been described previously, several combinations considered here have not been evaluated. We evaluated performance using Benjamini-Hochberg-adjusted P-values to account for multiple testing. Under the seven null scenarios, FWER was defined as the probability of at least one BH-significant cell type. With the exception of seismic, all methods adequately controlled FWER at 5% (**Fig. 2b**; results for individual null scenarios are shown in **Supplementary Fig. 1**), but differed substantially in power for detecting the causal β-cell type. MAGMA-GPA-based methods showed consistently lower power in these simulations than methods based on conLDSC or scDRS, despite generally maintaining FWER control. Among the individual methods, conLDSC-Cepo achieved the highest area under the receiver operating curve (AUC), demonstrating the benefit of using Cepo as a cell-type specificity metric within the conLDSC framework.

We next systematically searched for combinations of complementary methods to improve power while maintaining calibration. After excluding the two seismic methods that showed FWER inflation (**Fig. 2b; Supplementary Fig. 1**), we evaluated all equal-weight Cauchy combinations containing at least two of the 18 remaining methods, without requiring representation from each of the three methodological families (conLDSC, MAGMA-GPA and scDRS; **Methods**). We retained combinations for which the mean FWER below 5% across the seven null simulation scenarios. Among these well-calibrated combinations, we selected the smallest combination whose power was not significantly lower than the maximum observed power, based on leave-one-replicate-out comparisons across the four causal scenarios each configuration. This simulation-based procedure identified a parsimonious configuration, which we term CATCH (Cell-type Associations with Traits via CaucHy combination of strategies), combining conLDSC-Cepo and scDRS-mBAT-combo (**Fig. 2c,d**).

To investigate the robustness to incomplete scRNA-seq references, we reanalysed the simulated data using a four-cell-type endocrine-islet reference, either retaining or removing all β cells. The cell-type specificity statistics were recalculated using the remaining cells. Following β-cell removal, association signals across methods were consistently redistributed to transcriptionally related endocrine cell types, most frequently to δ cells, rather than disappearing completely (**Fig. 2e**). This result demonstrates that incomplete scRNA-seq references can redirect association signals toward biologically related cell types, highlighting the importance of reference completeness for accurate trait-cell type mapping.

### Establishing putative positive and putative negative trait-cell type pairs for benchmarking

We next constructed a literature-informed benchmark to enable method evaluation using real GWAS traits and scRNA-seq datasets (**Fig. 3a**). We selected GWAS summary statistics for 42 complex traits spanning 8 trait categories (**Supplementary Table 5**; genetic correlations among traits shown in **Supplementary Fig. 3**), each with ≥ 10 independent genome-wide significant loci (median=231) and SNP-based heritability > 0.05 (median=0.23). To systematically evaluate prior biological evidence for each trait-cell type relationship, we performed literature synthesis using 12 LLM configurations across different models and parameter settings (**Methods**)^16^. The prompts were designed to evaluate biological and mechanistic evidence independently of GWAS-based cell-type mapping studies, thereby minimizing information leakage from methods evaluated in this benchmark. We aggregated assessments across LLM configurations to reduce dependence on any individual model or setting. In parallel, we systematically searched PubMed titles and complete abstracts for evidence linking each trait and cell type, providing a transparent and reproducible measure of explicit literature support alongside the LLM-based synthesis.

**Figure 3.**
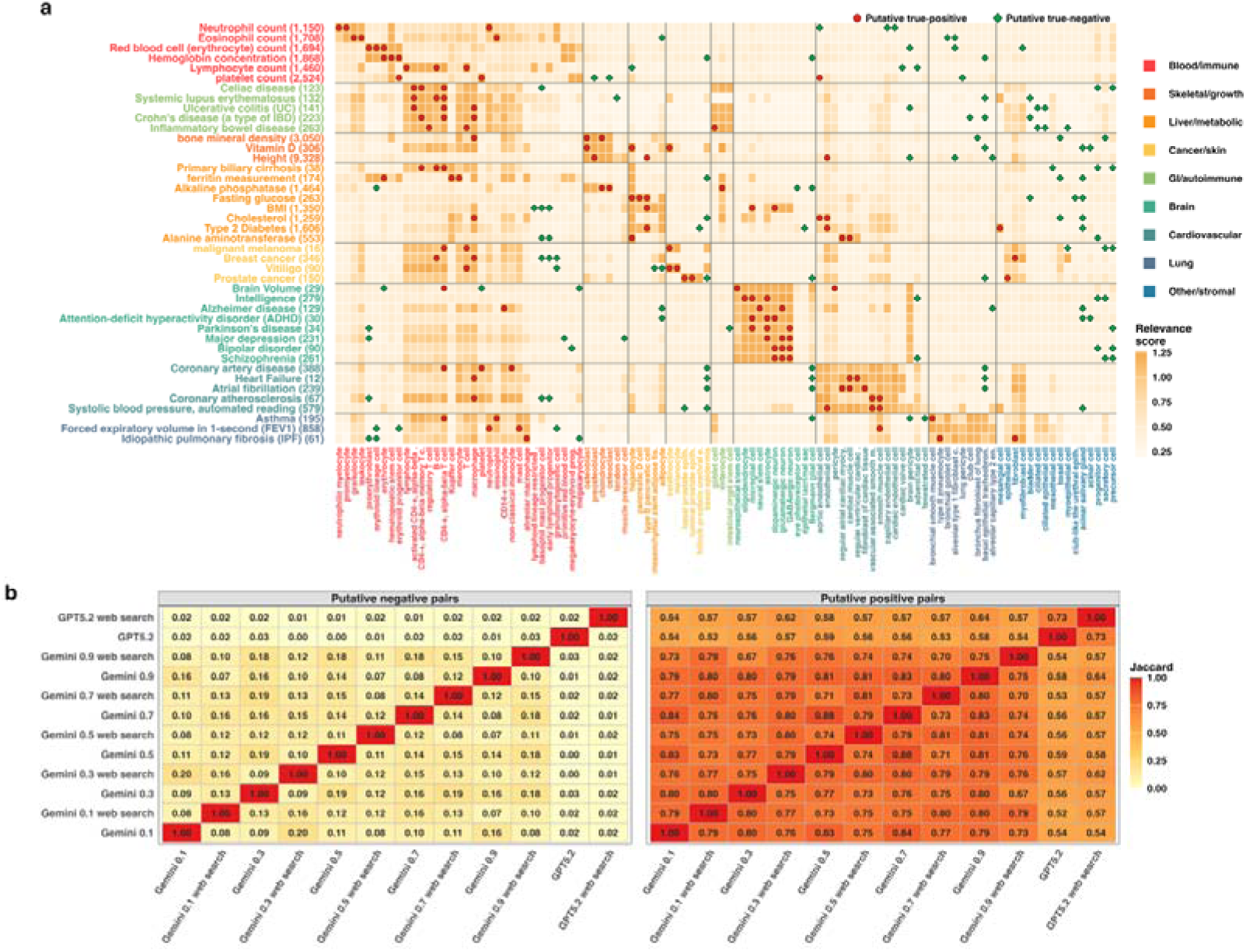
Construction and validation of an LLM- and PubMed-informed benchmark for trait-cell type mapping. **(a)** Literature-informed benchmark matrix including 42 complex traits and 104 cell types. Columns show cell types grouped by tissue (indicated by colour), and rows show complex traits and diseases, with the number of independent GWAS loci in parentheses. Traits are ordered and coloured corresponding to their relevant tissues. Colour intensity shows the literature-informed relevance score. Three putative positive cell types (red circle) and three putative negative cell types (green diamond) were selected for each trait based on their relevance scores, yielding 252 labelled pairs (126 putative positives and 126 putative negatives). Putative positive labels were defined as the top three top relevant cell types per trait, whereas putative negative labels were selected from low-scoring candidates using a minimum-overlap procedure to reduce repeated selection of broadly unrelated cell types. **(b)** Pairwise concordance of LLM-derived trait-cell labels. Heatmaps show the pairwise Jaccard similarity between LLM configurations for putative positive and putative negative labels across the benchmark trait-cell type pairs. Putative positive labels showed higher concordance than putative negative labels, reflecting consistent identification of biologically relevant associations while preserving diversity among putative negative labels.

We integrated these two measures of literature evidence into a continuous relevance score for each trait-cell type pair (**Methods**). Because publication frequency can reflect research intensity and historical hypotheses rather than biological relevance alone, PubMed evidence was given a secondary weight, with LLM consensus providing the primary contribution. We evaluated different PubMed weights and numbers of benchmark cell types, selected a PubMed weight of 0.25 and three high- and three low-relevance cell types per trait, which provided a balance between biological coverage and benchmark discrimination (**Supplementary Figs. 5 and 6**). The three high-relevance pairs and three low-relevance pairs were designated as putative positives and putative negatives, respectively, for each trait (**Fig. 3a**). These designations reflect current literature evidence rather than definitive biological truth, recognizing that biological knowledge is incomplete and evolving. Putative negative pairs therefore represent relationships with limited literature support rather than confirmed biological absence.

To assess the robustness of the benchmark, we evaluated the concordance of trait-cell type labels across 12 LLM configurations, including GPT-5.2 and Gemini with different temperature settings and search augmentation (**Methods**). Pairwise comparisons showed moderate to high concordance among LLM-derived putative positive labels (mean correlation r=0.71; **Fig. 3b**), indicating generally consistent identification of biologically relevant trait-cell type relationships. In contrast, putative negative labels showed low concordance (r=0.09), which was expected because putative negatives were selected from a broad set of low-relevance trait-cell type pairs. We consider this diversity advantageous because it avoids overcommitting to a single set of putative negative labels. By constructing benchmark labels from the LLM consensus scores rather than relying on any individual model, the benchmark is less likely to penalize methods that identify biologically plausible but previously underappreciated associations.

We further assessed whether differences in GWAS power across traits could bias the benchmark, particularly the putative negative pairs. Among the benchmark putative positive pairs, mapping signal increased modestly with the number of independent GWAS loci (r=0.28, P=0.010; **Supplementary Fig. 4**), consistent with greater statistical power to detect biologically relevant cell types in larger GWAS. In contrast, no significant association was observed for putative negative pairs (r=0.016, P=0.91). These results suggest that GWAS power primarily influences sensitivity to detect biologically relevant trait-cell type associations rather than the interpretation of putative negative benchmark pairs.

### Real-data benchmarking demonstrates superior performance of CATCH

We next evaluated whether the CATCH framework identified from the simulation study (**Fig. 2d**) generalized to real trait-cell type benchmarks derived from LLM- and PubMed-supported evidence (**Fig. 3**). The primary analysis used scRNA-seq data from the human atlas dataset (163 cell types) and 252 benchmark pairs across 42 GWAS traits, with three putative positive and three putative negative pairs for each trait (**Fig. 3a**). We compared all 21 benchmarked methods, including representative published methods and additional combinations generated from different cell-type specificity metrics and gene-based association methods **(Fig. 4a**; **Supplementary Table 3)**. Method significance was determined using Benjamini-Hochberg-adjusted P<0.05 within each method to account for multiple testing. Statistical power was evaluated as the proportion of putative positive pairs identified as significant, whereas empirical false discovery rate (FDR) was calculated as the proportion of significant benchmark pairs designated as putative negatives among all significant benchmark pairs.

**Figure 4.**
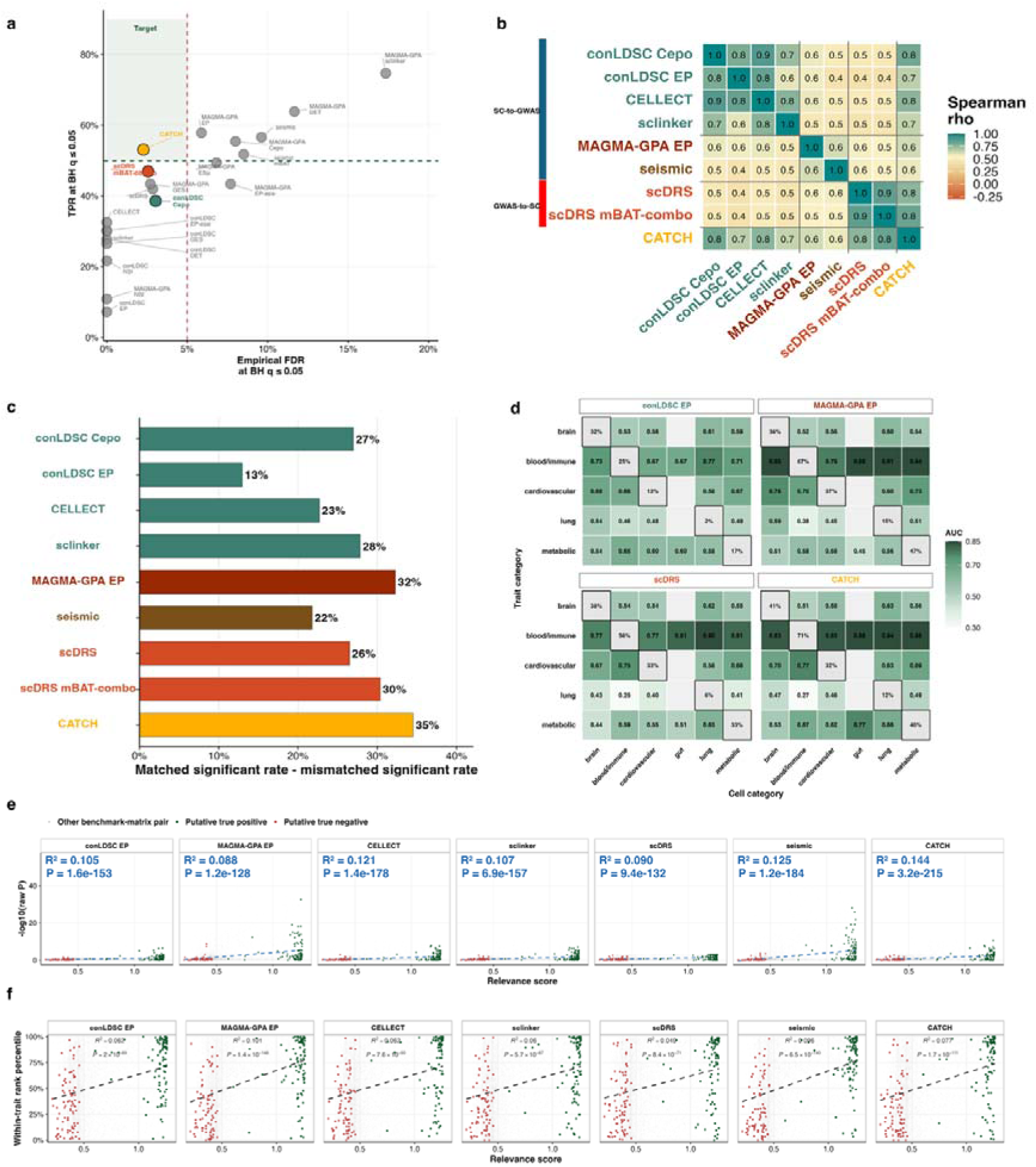
Real-data benchmarking demonstrates robust performance of CATCH. Putative positive and putative negative benchmark pairs are provided in Fig. 3. (**a)** Real-data benchmarking using the human atlas data. Statistical power (true positive rate, TPR) in y-axis and false discovery rate (FDR) at threshold of 5% in x-axis. Dashed lines mark the target performance criteria (power > 0.5 and FDR < 0.05). **(b)** Pairwise Spearman correlations between method-specific association statistics (-log10(P)) calculated across the same trait-cell pairs. **(c)** Bars show the matched significant rate minus the mismatched significant rate for each method. Larger values indicate that significant results are more enriched among biologically expected trait-cell pairs than among mismatched pairs. **(d)** Directional biological-system AUC across trait and cell categories. For each method, rows indicate the trait category, and columns indicate the cell category. Off-diagonal tiles show the AUC for separating biologically matched trait-cell pairs from mismatched pairs within the same trait category. Diagonal tiles show the percentage of matched trait-cell pairs with raw P < 0.05 and are outlined in black. Grey tiles indicate that the comparison was not available because no eligible matched or mismatched pairs were present. The analysis used six biological categories: brain, blood/immune, cardiovascular, gut, lung and metabolic, with 2,202 trait-cell pairs after pair-level averaging of duplicated resource rows, including 429 matched and 1,773 mismatched pairs. Gut is shown only as a cell-category column because no eligible gut-gut matched pairs were available after filtering. The complete comparison is shown in Supplementary Fig. 8. **(e)** Concordance between method association signals and the independently literature-derived relevance score across all benchmark trait-cell type pairs. Each point represents one trait-cell pair (Fig. 3a), with the literature-derived relevance score on the x-axis and method association strength (−log10(P)) on the y-axis. **(f)** Concordance between method prioritization ranks and the independently literature-derived relevance score across all benchmark trait–cell type pairs. Each point represents one trait–cell type pair (Fig. 3a), with the literature-derived relevance score on the x-axis and the within-trait method rank percentile on the y-axis (100% indicates the strongest association and 0% the weakest). Grey points denote other benchmark-matrix pairs, while green and red points denote putative true-positive and true-negative pairs, respectively.

As shown in **Fig. 4a**, conLDSC-EP, CELLECT, sc-linker and scDRS maintained low empirical FDR but also lower power. In contrast to its relatively low power in simulations, MAGMA-GPA-based methods showed higher power in the real-data benchmark, but this was accompanied by an elevated empirical FDR. Seismic-mBAT-combo and the standard seismic achieved higher power but exceeded the target empirical FDR. Consistent with the simulation results (**Fig. 2d**), CATCH achieved the strongest balance between sensitivity and empirical error control in the real-data benchmark (**Fig. 4a**). At method-wise BH P<0.05, CATCH detected 43 of 81 labelled positive pairs (power = 53.1%), with one false-positive calls among 44 labelled discoveries (empirical FDR = 2.3%). It was the only method to achieve a power above 50% while maintaining an empirical FDR below 5%.

To understand why CATCH outperformed individual methods, we compared method-specific association statistics (-log_10_(P)) across the same trait-cell type pairs. Methods within the same methodological family showed relatively strong concordance, whereas agreement between SC-to-GWAS and GWAS-to-SC approaches was only moderate (**Fig. 4b**; all pairwise comparisons are shown in **Supplementary Fig. 7**). This lower cross-family concordance indicates that the two strategies prioritize partly different trait-cell type associations, providing a rationale for combining complementary evidence across method families rather than relying on a single approach.

We next evaluated the robustness of the benchmarking results using two benchmark-label-free sensitivity analyses (**Methods**). Specifically, we (i) evaluated whether methods preferentially prioritized biologically matched systems, defined as trait-cell type pairs belonging to the same relevant biological system (for example, blood traits with immune cell types), over mismatched systems, and (ii) compared the continuous relevance scores with the association strength, measured by either -log_10_(P) or within-trait rank percentile, across all trait-cell type pairs. Across methods, biologically matched systems consistently showed stronger association signals than mismatched systems, with CATCH showing one of the largest within-versus-between system differences (**Fig. 4c,d**; **Supplementary Fig. 8**). Likewise, higher relevance scores were significantly associated with both stronger association signals and higher within-trait rank percentiles across all methods, where CATCH showed one of the strongest associations between relevance scores and both measures of association strength (**Fig. 4e,f**). Together, these benchmark-label-free analyses indicate that the superior performance of CATCH reflects biologically meaningful trait–cell type prioritization rather than dependence on predefined benchmark labels.

### Factors influencing genetic trait-cell type mapping performance

In the preceding sections we have provided simulation and real-data analyses to justify the CATCH strategy as the optimised method for declaring trait-cell type associations. Here, we investigate factors that influence the different methods proposed for GWAS and scRNA-seq integration.

#### Trait-cell type mapping depends on polygenic expression patterns rather than canonical marker genes

Both SC-to-GWAS and GWAS-to-SC strategies rely on gene sets. In the SC-to-GWAS strategy, these correspond to SEGs derived from cell-type-specific expression patterns, whereas GWAS-to-SC begins with a single set of trait-associated genes. We therefore first investigated how the choice of SEG metric influences downstream trait-cell type mapping. Using the human atlas dataset, we compared the top 10% of genes (a commonly used threshold^13,44^) across 8 commonly used cell-type specificity metrics (**Supplementary Table 2**) for three representative cell types (microglial cells, B cells, and pancreatic β-cells). The resulting gene sets overlapped by only about half (**Supplementary Fig. 9**), suggesting that different metrics capture distinct properties of the expression distribution of all genes within a cell type and of each gene across cell types. We then evaluated how well each metrics prioritize gold-standard marker genes used to provide cell-type labels across 28 cell types (≥ 5 marker genes per cell type). The differential expression t-statistic (DET) most accurately recovered canonical marker genes, followed by sc-linker^14^ and Cepo^28^ (**Fig. 5a**; **Supplementary Fig. 10**). However, this advantage did not fully translate into improved trait-cell type mapping. Instead, Cepo outperformed others in mapping power and empirical FDR control, regardless of whether GWAS enrichment was performed using conLDSC or MAGMA-GPA (**Fig. 5b**; corresponding FDR or false positive rate in **Supplementary Fig. 11**). One possible explanation is that canonical marker recovery emphasizes genes with the strongest differential expression, whereas genetic effects on complex traits are distributed across broader cell-type-specific transcriptional programs. Cepo, which prioritizes genes based on the consistency of their expression within a cell type relative to other cell types, may better capture such distributed signals. These results suggest that successful trait-cell type mapping depends on capturing polygenic cell-type-specific expression patterns rather than simply identifying canonical marker genes.

**Figure 5.**
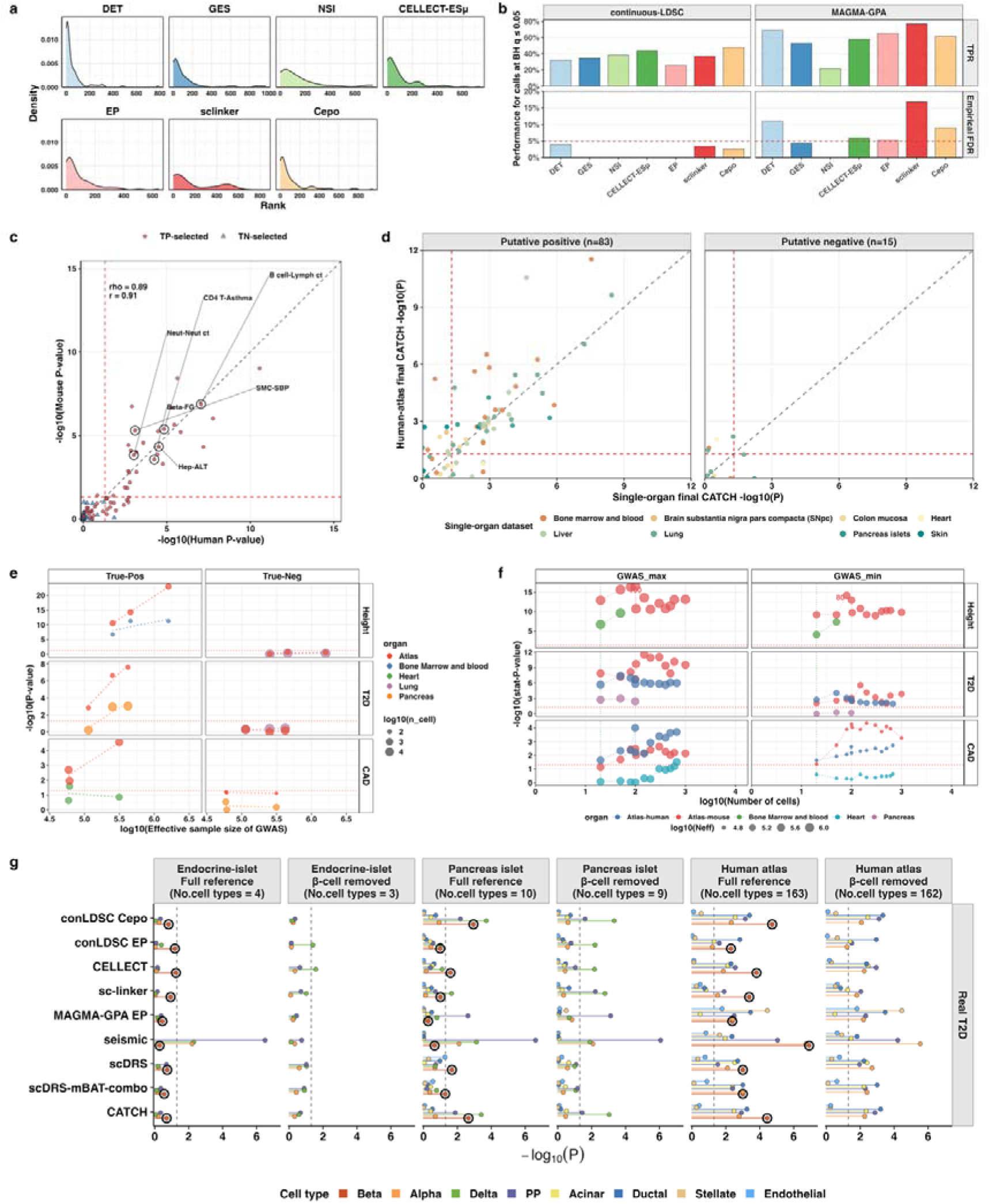
Factors influencing the performance of trait-cell type mapping methods. **(a)** Recovery of curated cell-type marker genes by different cell-type specificity metrics. Distributions of rank of cell-type specificity metrics show where gold-standard marker genes (**Supplementary Table 8**) rank under cell-type specificity metrics from DET, GES, NSI, CELLECT-ESμ, EP, sc-linker and Cepo (**Supplementary Table 2**). **(b)** Benchmark performance of individual cell-type prioritization metrics within the continuous-LDSC and MAGMA-GPA frameworks. Bars show median statistical power and FDR across putative positive and putative negative pairs. The dashed red line marks FDR = 0.05. Corresponding false positive rate is shown in Supplementary Fig. 11 **(c)** Cross-species reproducibility of CATCH signals using matched human and mouse atlas cell types. Each point represents one trait-cell type pair. Dashed lines indicate P = 0.05. Labelled examples were pre-specified representative putative positive and negative trait-cell type pairs spanning immune, lung, cardiovascular, pancreatic and liver-related biology. rho is the Spearman rank correlation between human-atlas and mouse-atlas CATCH results, calculated using the ranks of the putative positive and putative negative pairs. r is the Pearson correlation between the same human-atlas and mouse-atlas CATCH results (-log10(P)) for the same putative positive and putative negative trait-cell type pairs. (**d)** Comparison of atlas and single-organ references. Association strengths -log10(P) are compared between reference types for matched putative positive and putative negative trait-cell type pairs. The detailed complete comparison is shown in **Supplementary Fig. 19**. **(e)** Effect of GWAS power on trait-cell type association signals. Association strength (-log10(P)) is plotted against log10(effective GWAS sample size) for representative trait-cell type pairs for height, type 2 diabetes (T2D) and coronary artery disease (CAD). Point colour indicates dataset and point size indicates log10(number of cells). **(f)** Effect of target-cell number on association strength. Down-sampling curves show -log10(P) (y-axis) across increasing numbers of target cells (x-axis, minimum 20; log10(20) =1.3) for Height, T2D and CAD trait-cell type pairs. (**g)** Effect of removing β cells from the reference on T2D-cell type mapping across three reference contexts: a restricted endocrine-islet reference, a pancreas-islet reference and the full human atlas. Rows show individual methods and CATCH, and columns show each reference state, with the number of available cell types indicated in parentheses. Results are plotted for up to 9 relevant cell types, if available in the data set. Points show raw association strength (−log10(P)) for each displayed cell type. Horizontal line segments connect each point to zero to aid comparison within each method. Colours denote endocrine or islet-related cell types, and black outlines highlight β-cell results where β cells are present. The vertical dashed line indicates P = 0.05.

We also evaluated the choice of sLDSC significance statistics and SEG annotation strategies (**Methods**). Consistent with the recommendations of the sLDSC developers, τ P-values conditional on the baseline model (v1.2; 53 annotations^10^) showed better trait-cell type prioritization than heritability enrichment P-values (**Supplementary Figs. 12-14**). Compared with binary SEG annotations, continuous SEG annotations produced higher power in sLDSC and reduced FDR inflation in MAGMA-gene set analysis (**Supplementary Fig. 15**; **Supplementary Note**).

#### The impact of gene-based test choice depends on genetic architecture

We next investigated whether the gene-based test used to derive gene-level GWAS evidence affected trait-cell type mapping. Both scDRS and seismic depend on gene-level association statistics but aggregate them differently. We therefore compared MAGMA and mBAT-combo^45^ within otherwise matched scDRS and seismic configurations. We evaluated causal β-cell type recovery in the simulations and assessed power and FDR using human-atlas, mouse-atlas and single-organ benchmarks (**Supplementary Fig. 16**). In the causal simulations, mBAT-combo increased the β-cell association signal in both frameworks, with the largest improvement observed for seismic at *h*^2^=0.2. In contrast, scDRS and seismic showed similar power when using MAGMA or mBAT-combo in the real-data benchmarking, whereas differences in FDR control varied across reference resources. Thus, although mBAT-combo improved causal-signal detection in the simulations, particularly for seismic, it did not consistently improve trait-cell type mapping in real data. This implies that masked genetic associations (where causal variants have negative genetic covariance), for which mBAT-combo is better powered to detect, are not an important characteristic of the genetic architecture of the GWAS traits studied here; this may need to be re-evaluated in future as GWAS become even more powered^39^.

Overall, genes that featured by SC-to-GWAS and GWAS-to-SC approaches were enriched for disease- and cell-type-relevant functions, whereas genes unique to each strategy captured broader, complementary programs related to cell identity and genetic disease association, respectively. For instance, functional enrichment of T2D pancreatic β-cell genes showed that genes prioritized by both Cepo and MAGMA were enriched for diabetes- and β-cell-related pathways. Cepo-only and MAGMA-only genes captured broader, complementary functions related to pancreatic cell identity and insulin secretion, and cell-cycle and disease-associated processes, respectively (**Supplementary Fig. 17**).

#### Human and mouse scRNA-seq references produce largely consistent trait-cell type mapping

To investigate the impact of the choice of species from which scRNA-seq references were generated, we focused on matched cell types from a human atlas^39^ and a mouse atlas^46^ dataset generated by the same lab, with all cell types available used to generate SEGs. Although the human atlas contained substantially more cells than the matched mouse atlas (**Supplementary Fig. 18**), CATCH clearly distinguished putative positive from putative negative benchmark pairs in both species (**Fig. 5c**). Across matched trait-cell type pairs, association signals were highly concordant between human and mouse references (r = 0.796, P = 2.0 × 10^−1^^8^), with no significant difference in mean association strength (paired *t*-test P = 0.26). These findings indicate that human and mouse scRNA-seq references produce broadly consistent trait-cell type mapping results, consistent with previous findings^25^ and supporting the use of mouse datasets for initial discovery when human data are unavailable.

#### Diverse atlas-scale references improve mapping power

Although scRNA-seq datasets are increasingly available, most have been collected from a single organ due to cost, complexity, and specific research interests. To assess whether using a single-organ dataset affects the conclusions about method performance, we repeated our analyses across datasets from different organs, while ensuring comparable pairwise evaluations to those based on atlas datasets. Notably, atlas-scale scRNA-seq references consistently outperformed single-organ datasets in power using CATCH (**Fig. 5d**) as well as across both SC-to-GWAS and GWAS-to-SC methods (**Supplementary Fig. 19a**). All methods could fail to detect some literature-supported trait–cell type (e.g. IBD-goblet cells^47^), which may indicate the relationships between genetic regulatory effects and cell-type transcriptional programs depending on inflammatory stimulation, environmental exposure, or disease state etc. Consequently, reference profiles obtained from healthy or resting cells may not capture the context-specific gene programs through which these cell types contribute to disease. Meanwhile, FDR were effectively controlled in the putative negative pairs (**Supplementary Fig. 19b**). For the SC-to-GWAS strategy, the greater diversity of comparator cell types produced more stable estimates of cell-type specificity (**Supplementary Fig. 20**). For the GWAS-to-SC strategy, broader references improved the calibration of disease and control scores across cells, resulting in stronger and more reproducible trait-cell type associations (**Supplementary Figs. 21-25; Supplementary Note**).

#### GWAS power and cell type diversity jointly determine mapping performance

Having established the importance of reference diversity, we next evaluated the relative contributions of GWAS sample size and the number of cells per cell type in the scRNA-seq data. Using height, type 2 diabetes (T2D), and coronary artery disease (CAD), for which GWAS of different samples sizes are available, we found that increasing GWAS sample size consistently strengthened trait-cell type associations while maintaining FDR control across methods (**Fig. 5e**). In contrast, increasing the number of cells per cell type produced diminishing returns, with statistical power plateauing once approximately 80-150 quality-controlled cells were available for well-powered traits such as height and T2D (**Fig. 5f**; **Supplementary Fig. 26; Supplementary Note**). As most cell types in both the human and mouse atlas datasets already exceeded this threshold, further improvements in trait-cell type mapping are likely to arise primarily from larger GWAS and more comprehensive reference atlases rather than sequencing additional cells within existing cell types.

#### Reference diversity improves robustness when causal cell types are missing

Because no scRNA-seq reference captures every biologically relevant cell type, we next asked whether broader cell-type diversity helps stabilize trait-cell type mapping when an expected causal cell type is absent. Using the same real T2D GWAS summary statistics, we compared three real data based single cell reference contexts with increasing cellular diversity: a restricted endocrine-islet reference (4 cell types), extracted subset of the full pancreas-islet (10 cell types), the full pancreas-islet reference and the full human atlas (163 cell types) to evaluate the robustness of the results generated by different methods before and after removing the causal cell type that we believed, pancreatic β cells in these three reference datasets.

In the restricted endocrine-islet reference, removing β cells redistributed association signals among the three remaining endocrine cell types (**Fig. 5g**). Several methods shifted the trait association signal toward δ cells, whereas seismic-based methods retained stronger pancreatic polypeptide cells (PP cells) signals, though this association was no longer significant. This indicated that, in a constrained reference, removal of the expected causal cell type can substantially change the apparent top-ranked cell type. In contrast, the broader pancreas-islet and human-atlas references showed greater stability. In the human atlas, CATCH retained strong signals after β-cell removal. These results suggest that broader cell-type diversity provides additional biological and transcriptional context, reducing the extent to which the result depends on one available cell label. Thus, atlas-scale or otherwise diverse reference datasets can preserve more interpretable trait-cell association signals even when an expected causal cell type is missing. The variation in significant trait–cell type signals across reference datasets further highlights the importance of background cell-type composition for robust trait–cell type mapping.

#### Using only protein-coding genes is sufficient for trait-cell type mapping

In addition to cell types, the selection of background genes may also affect trait-cell type mapping. Most studies focus on protein-coding genes for their direct functional relevance relative to other gene biotypes (e.g., long noncoding RNAs, short noncoding RNAs and micro RNAs^48^), although an unknown fraction of non-coding genes may contribute to complex trait variation^49,50^. We therefore compared mapping performance of all expressed genes versus only protein-coding genes in both atlas and single-organ scRNA-seq datasets. Despite substantial variation in the number of expressed genes across organs, restricting analyses to protein-coding genes produced comparable performance to using all expressed genes (**Supplementary Fig. 27**). These results suggest that protein-coding genes capture most of the information required for current trait-cell type mapping analyses. However, this may partly reflect that protein-coding genes are more likely to have well-established functional roles and are more comprehensively characterized than non-coding genes, rather than an absence of contributions from non-coding genes.

### A practical decision framework and relevance resource for trait-cell type mapping

Finally, we translated the benchmarking results into a practical decision framework for researchers applying trait-cell type mapping methods to new traits or diseases (**Fig. 6**). The framework supports two common use cases. For method developers and benchmarking studies, we provide a curated relevance resource together with putative positive and putative negative trait-cell type pairs. This will enable evaluation of future methods against the benchmark established in this study. For researchers applying trait-cell type mapping to new traits or diseases, the framework guides method selection according to the available data, including GWAS summary statistics, individual-level scRNA-seq data, cell-type specificity scores, or SNP-level genomic annotations. The recommended workflow prioritizes CATCH, which integrates conLDSC-Cepo and scDRS-mBAT-combo, when the inputs required for both components are available, and provides alternative analyses when only partial data are available. By making the data requirements and analytical assumptions explicit, this framework provides a practical and reproducible route from benchmarked methodology to biological discovery.

**Figure 6.**
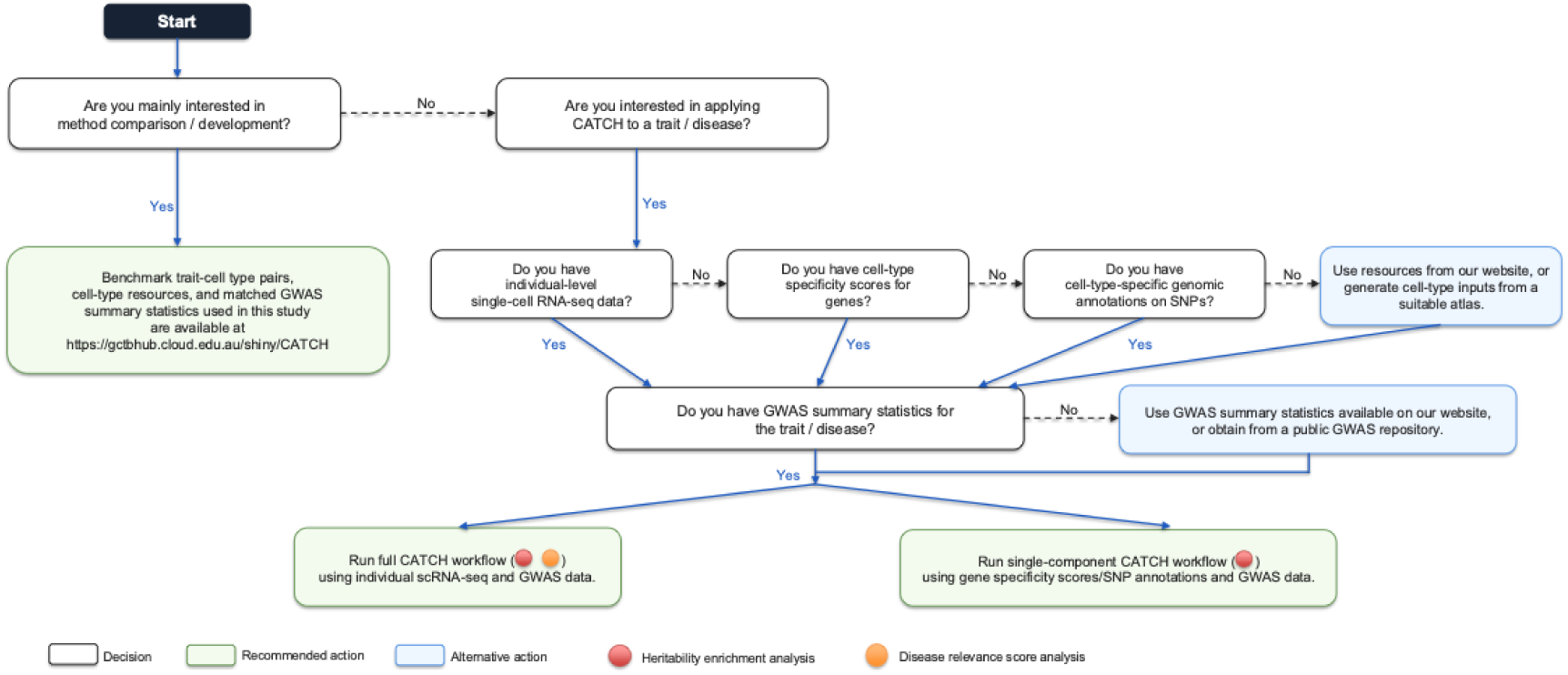
Practical guidelines and relevance resource for trait-cell type mapping. For method development, the framework provides a literature-informed benchmark comprising curated trait–cell type benchmark pairs and relevance scores, enabling standardized evaluation of new methods. To apply CATCH to new traits or diseases, complementary trait-cell type mapping methods are combined according to the available input data, including GWAS summary statistics, individual-level scRNA-seq data, cell-type specificity scores, or SNP-level genomic annotations.

## Discussion

In this study, we evaluated methods for identification of cell types for complex traits under the hypothesis that trait-associated genes will show gene expression specificity in trait-relevant cell types (**Fig. 1**). We developed a comprehensive benchmarking framework that integrates controlled simulations (**Fig. 2**), an LLM-assisted literature-informed benchmark (**Fig. 3**), and multiple benchmark sensitivity analyses. This framework enables a systematic comparison of existing methods and led to the development of CATCH, which consistently increased power of discovery while maintaining false-positive control across diverse benchmark settings (**Figs. 2** and **4**).

Our results also provided several insights into the design of trait cell-type mapping methods. First, identifying canonical marker gene is not sufficient for accurate trait-cell type mapping. Although DET most effectively recovered curated marker genes used for cell-type annotation, Cepo^28^ provided superior performance for mapping trait-associated cell types (**Fig. 5a, b**). This distinction is biologically intuitive because marker genes are optimized to distinguish cell identities^25^, whereas polygenic effects underlying complex traits are distributed across many genes that may be shared across multiple cell types. Consequently, methods that capture broader patterns of cell-type-specific expression outperform those optimized solely for marker-gene detection. Second, neither the SC-to-GWAS nor the GWAS-to-SC strategy identified all putative positive trait-cell type pairs. Instead, the moderate concordance observed between the two methodological strategies indicates that they capture complementary rather than redundant biological information (**Fig. 4b**). This complementarity provides a principled explanation for why combining evidence across method families improves robustness and underlies the performance of CATCH.

The final CATCH configuration included conLDSC-Cepo and scDRS-mBAT-combo but not a MAGMA-GPA component. In simulations, adding MAGMA-GPA provided little incremental information beyond these two complementary approaches. In the real-data benchmark, MAGMA-GPA showed higher power but also identified more putative negative pairs. This difference may reflect the greater complexity of genetic architectures in real traits and the imperfect correspondence between literature-informed benchmark labels and biological relevance, such that some additional MAGMA-GPA discoveries may represent associations not captured by current literature. We therefore adopted a conservative strategy and excluded MAGMA-GPA from CATCH to prioritize robust error control.

Our benchmarking also provides practical guidance for future studies. Among the SC-to-GWAS methods evaluated, continuous cell-type-specific annotations generally outperformed binary annotations (**Supplementary Fig. 15**), and sLDSC performed the best with Cepo, whereas MAGMA-GPA performed the best with EP (**Fig. 5b**). Discovery of trait-cell type association is comparable when using scRNA-seq data derived from either mouse or human origins (**Fig. 5c**). Increasing GWAS sample size substantially improved trait-cell type mapping power, whereas increasing the number of cells per cell type showed diminishing returns once approximately 80-150 high-quality cells were available for well-powered traits (**Fig. 5d, e**). These observations suggest that future improvements in trait-cell type mapping are likely to arise primarily from increasingly powerful GWAS rather than from substantially larger single-cell datasets for the known cell types.

We further found that atlas-scale scRNA-seq references or single-organ datasets with high cellular diversity yielded higher power than single-organ datasets with limited cell-type representation and produced more stable trait-cell type mapping when expected causal cell types were absent (**Fig. 5f, g**). These findings emphasize that the value of a reference dataset depends not only on whether it contains the nominally relevant tissue but also on the diversity of biologically related cell types available for comparison. In practice, one should consider whether a reference captures the breadth of plausible disease-relevant cell types, especially for systemic traits, immune-mediated diseases or traits with uncertain tissue origins. From the perspective of single-cell datasets, more emphasis should be placed on increasing cell-type diversity within the datasets themselves. This depends not only on developing or improving technologies and methods to resolve novel cell types and increase resolution, but also on expanding the range of conditions sampled, e.g. disease context, stimulation, and developmental timepoints, to capture a broader spectrum of cell states. Potential improvements to methods such as scDRS, for example by selecting control genes from the same cell type rather than from the whole background when analysing rare cell types.

Our study also highlights the value of a literature-informed benchmark. Rather than relying solely on publication counts, we integrated evidence across multiple LLMs together with PubMed while explicitly reducing the influence of publication frequency. We provide LLM prompts for researchers to keep up date if they interest in the latest LLMs considering they are fast-moving. High concordance among putative positive labels indicated consistent recognition of biologically relevant trait-cell type relationships, whereas the deliberate diversity among putative negative labels reduced reliance on any single definition of biological irrelevance (**Fig. 3**). The benchmark-label-free analyses further demonstrated that the conclusions did not depend on predefined binary benchmark labels, providing additional confidence in the robustness of the evaluation framework. To improve the biological interpretation of GWAS signals, an increasing number of methods integrate genetic associations with modalities beyond scRNA-seq, including spatial transcriptomics and single-cell chromatin accessibility data. Examples include gsMap^51^, which maps trait associations using spatial transcriptomics, and SCAVENGE^52^, which identifies trait-relevant cells from single-cell chromatin accessibility profiles. Beyond benchmarking GWAS–scRNA-seq integration methods, our curated trait-cell type benchmark could provide a reusable reference for evaluating methods across diverse single-cell and spatial-omics modalities. This broader applicability highlights systematic multimodal benchmarking as an important direction for future research.

Several limitations should be considered. First, although the literature-informed benchmark substantially improves over simple publication-count approaches, it remains limited by current biological knowledge and cannot capture novel trait-cell type relationships that have yet to be described. Moreover, trait-cell type associations inferred from GWAS and expression specificity do not by themselves establish causal cellular mechanisms. They should be interpreted as prioritizations that require follow-up using functional genomics, perturbation experiments or disease-relevant cellular models. Second, although our simulations used real genotype data and a human atlas scRNA-seq dataset, the simulated genetic architectures might simplify the complexity of real traits. We considered multiple causal and null scenarios, but real traits may involve multiple relevant cell types and more complex genetic architectures. Nevertheless, the conclusions from the simulations were independently supported by the literature-informed benchmark across 42 real traits and multiple atlas- and organ-scale scRNA-seq references, indicating that the overall findings are not dependent on the simulation design. Third, our benchmark focused primarily on broad cell types and therefore did not assess performance at the level of individual cells or fine-grained cellular states. This limitation is particularly relevant to scDRS, whose ability to prioritize individual cells without predefined cell-type or cluster annotations distinguishes it from the other evaluated methods. This capability may reveal disease-relevant cellular states or subpopulations that are obscured by broader cell-type classifications. Extending the benchmark to single-cell resolution will require independent ground truth at the cellular-state level, which is considerably more difficult to establish than trait-cell type relationships. Developing such benchmarks is therefore an important direction for future research. Fourth, the benchmark was constrained by the availability of sufficiently powered GWASs and corresponding cell types in scRNA-seq datasets. Nonetheless, the robustness of results using single cell data from model species is encouraging given that some environmental exposures relevant for diseases are much more easily explored using non-human species. Fifth, beyond these data-coverage constraints, benchmark interpretation also depends on the validity of the reference labels. For example, two putative negative endothelial pairs for the trait of neutrophil counts produced strong CATCH signals (**Supplementary Fig. 19**). These discordant signals highlight the inherent uncertainty of constructing negative reference pairs for trait-cell type associations. The putative negative labels reflect limited prior evidence rather than evidence that an association is biologically absent. The endothelial signals may reflect incomplete literature coverage, shared vascular gene programmes, or nonspecific enrichment arising from overlapping gene sets. Therefore, these pairs should not be interpreted as definitive false positives or evidence of systematic inflation. Sixth, differences between scRNA-seq and snRNA-seq are not considered. snRNA-seq may miss important gene expression signals^12^, such as microglial activation shown by previous research^53^. However, our study was limited to available data, and technical limitations mean that only snRNA-seq data can be generated for some organs, such as brain. Seventh, part of the differences between single-organ and atlas datasets may be due to technical aspects (data generated by different labs) which is out of our control. A completely fair comparison is difficult, although we did subset atlas data to make single organ data sets. Eighth, we focused only on the methods using scRNA-seq and GWAS data with common SNPs. Methods incorporating additional information, such as rare variants (EPIC^17^), QTLs (ECLIPSER^54^), or tissue enhancer annotations (sc-linker^14^), were beyond the scope of this study but are expected to further improve trait-cell type mapping^2,55,56^. Likewise, methods developed for different objectives, such as TWAS-based approaches (e.g., scPrediXcan^57^) or prediction of cell-type-specific gene expression (e.g., ctPred^58^), were not included because they address distinct biological questions.

Overall, our study provides a reproducible benchmarking framework, a curated trait-cell type relevance resource, and practical recommendations for future development and application of trait-cell type mapping methods. As GWAS continue to increase in statistical power and increasingly comprehensive single-cell atlases become available, this framework can be readily extended to benchmark emerging methods and additional molecular modalities, facilitating increasingly accurate identification of disease-relevant cell types.

## Online Methods

### Benchmark construction

#### LLM-assisted relevance assessment

Trait-cell type relevance was evaluated using 12 LLM configurations accessed through their respective Application Programming Interfaces (APIs). These comprised GPT-5.2 with and without web search and Gemini configurations across five temperature settings (0.1, 0.3, 0.5, 0.7 and 0.9), each with and without web search, to assess the robustness of trait–cell type relevance rankings to stochastic variation in LLM outputs. For each trait and cell type included in this study, a standardized prompt (**Supplementary Data 1**) was submitted using a Python workflow that generated the prompt, executed the model in an isolated environment, and stored the structured output. The prompts explicitly instructed the models to evaluate trait-cell type relevance based on biological and mechanistic evidence while excluding evidence from GWAS-based cell-type mapping studies and the methods evaluated in this benchmark, thereby minimizing information leakage. All analyses were performed using Python 3.11.14 and Node.js, and the scripts and prompts are available at **Supplementary Note**.

#### LLM-derived relevance scores

For each trait-cell type pair (*t*, *c*), each LLM configuration *m* returned a raw relevance score *r*_tcm_. Because these scores were not directly comparable across traits or models, they were converted into within-trait empirical percentiles,

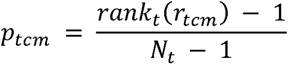

where *N*_t_ denotes the number of candidate cell types for trait *t*. The quantity *p*_tcm_ therefore lies on the interval [0,1], with larger values indicating that cell type *c* was ranked as more relevant to trait *t* by model *m*.

For each trait-cell pair, we summarized the 12 model-specific percentiles by three statistics. First, we defined the mean percentile across models:

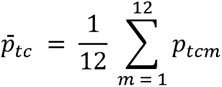

This term captures the average relative relevance of the pair across all 12 LLM configurations. Second, we counted how many models placed the pair in the top 20% of cell types for the same trait:

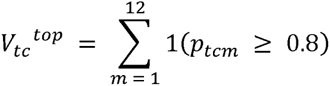

where *1*(.) is the indicator function. *V*^top^_tc_ measures repeated high-confidence support across models. Third, we counted how many models placed the pair in the bottom 20% of cell types for that trait:

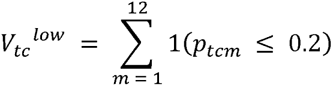

*V*^low^_tc_ therefore measures repeated low-relevance support across models. We combined these three quantities into a consensus relevance score:

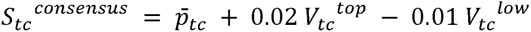

In this construction, the mean percentile provided the primary ranking, whereas the vote terms acted only as tie-breakers among similarly ranked pairs.

#### PubMed scores

For each trait–cell type pair, we obtained the number of PubMed publications matching the trait and candidate cell type. Publication counts were log-transformed and converted to within-trait empirical percentiles, allowing direct comparison across cell types while reducing the influence of highly studied traits and cell types. These percentile scores were subsequently incorporated into the literature-informed relevance score.

#### LLM- and PubMed-weighted consensus relevance scores

The final relevance score combined the within-trait percentile of the 12-model consensus relevance score *p_tc_*^(cons)^ with the within-trait percentile of log-transformed PubMed support *p_tc_*^(PubMed)^:

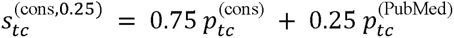

To evaluate the influence of literature support, we constructed benchmark scores using multiple PubMed weights (0, 0.25, 0.5, 0.75 and 1.0), where a weight of 0 represents an LLM-only benchmark and a weight of 1.0 corresponds to a PubMed-only benchmark. The benchmark used throughout the main analyses employed a PubMed weight of 0.25. The rationale for this choice and the corresponding sensitivity analyses are presented in **Supplementary Fig. 6**.

#### Construction of benchmark trait-cell pairs

For each trait, candidate pairs were ranked using the weighted consensus relevance score, *S_tc_*^(cons,0.25)^. Putative positive pairs were selected from the highest-ranked eligible cell types. Putative negative pairs were selected from low-ranked candidates after excluding generic labels, immune-cell negatives, PubMed-supported pairs and cell types within the dominant relevant organ or cell-category context for that trait. When the fully constrained negative pool was too small, organ separation was relaxed before cell-category separation, while the non-generic, non-immune and PubMed-unsupported requirements were retained. This design contributed three putative positive and three putative negative labels per trait across 42 traits shown in **Figure 3a**.

#### Benchmark-label-free analyses

To evaluate methods without relying on predefined binary benchmark labels, we performed two complementary analyses. First, traits and cell types were grouped into broad biological systems, and methods were evaluated according to the difference in mean association signal between biologically matched and mismatched systems (Cardiovascular, Blood, Brain, Immune/GI, Lung, and Metabolic/Liver). Second, continuous literature-derived relevance scores were compared directly with method-specific association statistics across all available trait-cell type pairs.

#### Simulation

We performed simulations using HapMap3-matched genotypes from 20,000 unrelated UK Biobank participants of inferred European ancestry. Pancreatic β cells were specified as the causal cell type using an externally curated set of 94 β-cell marker genes. We simulated four β-cell-enriched causal scenarios by crossing two genetic architectures with SNP heritability of h^2^=0.1 and 0.2. Each scenario contained 10,000 causal HapMap3 variants, of which either 2,000 (20%) or 5,000 (50%) mapped within 100□kb of β-cell-specific genes; the remaining variants were sampled from the non-overlapping genomic background. We also evaluated seven null scenarios: a no-effect model (h^2^=0); genome-wide random, functional-annotation-enriched and infinitesimal architectures, each simulated at h^2^=0.1 and 0.2. None of the null architectures incorporated cell-type-specific annotations. Each causal and null scenario comprised 30 replicates. Phenotypes were simulated using GCTA and analysed using fastGWA to generate GWAS summary statistics. Atlas-scale trait–cell-type mapping was performed using 155 cell types from the human cell atlas reference (155 out of 163 cell types detected by all methods). For the β-cell-removal sensitivity analysis, we used a separate endocrine-islet simulation dataset comprising ten replicates at each heritability level. The h^2^=0.1 simulations contained 800 β-cell-gene-mapped and 200 background causal variants, whereas the h^2^=0.2 simulations contained 900 and 100 variants, respectively. Each method and the CATCH configuration were evaluated using both the complete four-cell endocrine-islet reference and the corresponding three-cell reference obtained by removing β cells.

#### Simulation-based selection of CATCH

CATCH was selected using simulation results only. We evaluated all equal-weight Cauchy combinations containing at least two of the 18 non-SEISMIC method outputs, without requiring representation from each method family, yielding 262,125 candidate combinations = 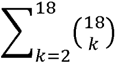 = 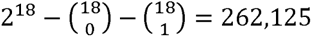. For each combination, we calculated the mean family-wise error rate (FWER) across the seven null scenarios and the maximum scenario-specific FWER. We retained combinations for which both measures were below 5%. Among these well-calibrated combinations, the configuration with the highest mean β-cell detection power across the four causal scenarios was used as the reference. We then identified configurations whose power was not significantly lower than that of the reference using leave-one-replicate-out jackknife estimates and a one-sided lower-tail t-test with 29 degrees of freedom (30 replicates for each scenario). Among these configurations, we selected the one containing the fewest component methods. This procedure selected the equal-weight combination of conLDSC-Cepo and scDRS-mBAT-combo as the final CATCH configuration.

#### Controlled single-cell reference datasets

We evaluated simulations and reference-perturbation analyses using three scRNA-seq reference families. First, simulations were performed using a reduced endocrine-islet reference derived from the Fasolino et al.^40^, comprising the four principal endocrine cell types (β, α, δ and PP cells). A matched β-cell-removed reference was generated by removing all β cells to assess the impact of missing causal cell types in a constrained endocrine context. Second, we used the full Fasolino pancreas reference to evaluate β-cell removal in a broader pancreatic cellular context while preserving both endocrine and exocrine cell populations. Third, we used the Tabula Sapiens Human Cell Atlas to evaluate β-cell removal in a large multi-organ reference. A matched β-cell-removed atlas was generated by removing all pancreatic β cells. Together, these three reference datasets allowed us to evaluate (i) controlled causal-cell recovery in a restricted endocrine reference, (ii) robustness in a pancreas-scale reference, and (iii) generalizability in a diverse multi-organ atlas.

#### scRNA-seq data pre-processing

We used 10 databases of scRNA-seq data (**Supplementary Table 3**). Considering the high level of conservation with one-to-one orthologous relationships for protein-coding genes between mice and humans, we focused on 19,430 such genes with 19,116 expressed in the human (TS) atlas data and 14,879 expressed in the mouse (TMS) atlas data for the analysis. We standardized the cell-by-gene matrix by reversing prior log normalization to ensure dataset consistency, followed by standard UMI-based normalization to adjust for sequencing depth variations. The final expression values are represented as log2(TPM + 1), where TPM is transcripts per million. Cell type annotations were initially based on existing labels in the dataset provided by the group generating and releasing the data. We then manually standardized these labels to ensure identical naming for the same cell types, correcting any inconsistencies in the original annotations. For cross-species analysis, mouse genes were mapped to their orthologous human counterparts using Ensembl (v. 91), with only one-to-one mapping orthologs retained.

#### GWAS data QC

We performed PLINK clumping to calculate the number of independent loci (--clump-p1 5e-8 -- clump-r2 0.1 --clump-kb 500) and SBayesRC (GCTB_v2.5.2) to estimate the SNP-based heritability (on liability scale) for a range of complex traits and diseases. We selected 42 complex traits and diseases for which 42 GWAS summary data with a large sample size (n >13,000) are publicly available and have a polygenic architecture (number of independent GWAS loci > 10 and SNP-based heritability > 0.05). The trait information can be found in **Supplementary Table 5**.

### Identifying associations between traits/diseases and cell types

#### Calculation of cell-type specificity metrics

The SC-to-GWAS strategy usually includes two steps. The first step is to identify the cell type specifically/differentially expressed genes or cell identity genes. Here, we used SEG (specifically expressed genes) to represent these genes. Briefly, predefined cell type labels are required, and a cell type label is usually based on very high gene expression of a small number of genes. Various statistical methods are available to calculate the cell type specificity score **Supplementary Table 2**. In each method genes in the top decile (i.e., 1,400 genes) are considered as SEGs (e.g., TDEP, top decile expression proportion) and are used to create either a binary genomic annotation (e.g. LDSC-SEG and MAGMA covariate analysis, also refers to MAGMA-gene-set-enrichment analysis or MAGMA-GSEA) or a continuous genomic annotation (e.g. sc-linker, CELLECT and MAGMA gene property analysis) for each cell type. We ran the following cell-type specificity metric methods using their default parameter settings: EWCE^27^ for EP, Cepo^28^, CELLEX^13^, and sc-linker^14^. All methods followed the same pre-processing steps for scRNA-seq data, as described above. Furthermore, CELLEX filtered out sporadically expressed genes using a one-way ANOVA with cell type annotations as the grouping factor. EP and CELLEX do not have the requirement for the minimal number of cells within each cell type. In contrast, scDRS includes cell types with more than 3 cells, sc-linker includes those with more than 10 cells and Cepo includes those with more than 20 cells. Although different metrics have their own criteria for the minimal number of cells within the cell type, we excluded cell types with fewer than 20 cells in our analysis.

#### GWAS and scRNA-seq integrative analysis

The GWAS-to-SC strategy requires the results of gene-based test using GWAS data as input for the integrative analysis, such as scDRS. We compared the performance of scDRS using MAGMA (as used in the scDRS publication) and mBAT-combo (a new method which is better powered for gene associations when haplotypes have risk alleles with opposite effect) as the gene-based test methods, respectively. For a fair comparison, we used the transcription start and end site +/- 10kb window (suggested by scDRS default settings) to define a gene region when conducting gene-based tests using MAGMA (v1.09) and mBAT-combo. The 1000 Genomes Project phase 3 European genotypes were used as the LD reference data.

For the SC-to-GWAS strategy, we used sLDSC and MAGMA gene-set enrichment methods. sLDSC tests whether the 10% most specifically expressed genes in a given cell type (define from specificity metrics) are enriched in SNP-based heritability for the trait. Only genes with at least 1 TPM or 1□UMI per million in the tested cell type were used for this analysis. To capture potential regulatory elements contributing to the effect of the region on the trait, we extended the gene coordinates by 100□kb upstream and downstream, respectively, for each gene (suggested by all methods based on the sLDSC framework). SNPs located in 100 kb regions surrounding the 10% most specific genes in each cell type were annotated with either binary or continuous values. This cell type-specific genomic annotation was analysed in sLDSC for one cell type at a time, jointly with 53 functional genomic annotations included in the baseline model v1.2^44^. The coefficient z-score P value was used as a measure of the association of the cell type with the trait. The significance threshold was set to be 5% empirical FDR across all tissues/cell types and traits within each dataset. We also evaluated the cell-type specificity gene sets in the MAGMA-GSEA approach. In this approach, we examined the association between MAGMA gene-level trait associate statistics and different SEG metrics with either binary or continuous values using MAGMA-GPA test. We adjusted the gene-level z-score obtained from MAGMA for default covariates, including gene size, gene density (a measure of within-gene LD), inverse mean minor allele count, as well as the logarithm of these variables. Subsequently, we performed a linear regression model, with the MAGMA gene-level z-score as the dependent variable and SEG metrics for each cell type as the independent variable. The outcome of interest was the significance of the contribution of cell type SEG to the regression coefficient of gene-level z-scores of trait association, assessed through a one-sided test (whether the gene-level z-score is positively associated with the SEG metric). In summary, this analysis allowed us to calculate cell type prioritization P-value, indicating the positive impact of SEG on the trait’s gene-level z-statistics, as determined by the linear regression model. In the structure of *Seismic*, the disease genes z-statistics could be replaced with mBAT-combo based z-statistics to become Seismic-mBAT-combo.

## Acknowledgements

We gratefully acknowledge DPhil Malte D. Luecken, A/Prof. Nathan Palphant and Dr. Sonia Shah for consultation on true-positive pairs selection in relative fields. This research was funded by the Australian National Health and Medical Research Council (1173790, 1177268, 2043111), the National Institute for Mental Health (1RO1MH124871-01), Oxford Health BRC (NIHR203316) and the University of Oxford Michael Davys Trust. J.Z. acknowledges support from Australian Research Council grant FL180100072 (awarded to P.M. Visscher). This study makes use of data from UK Biobank (project ID: 116122).

## Contributions

Conceptualization: A.L., J.Z., N.R.W., Methodology: A.L., J.Z., N.R.W., Formal analysis and investigation: A.L., GWAS data QC and format, SNP-heritability on liability scale estimates: T.L., S.W. Genetic correlation estimate: S.W., Independent number of SNPs estimates: R.Zh. S.W, Cell ontology terms match: A.L., Y.S, R.Zh., LLM prompt and script: H.C., binary-LDSC: A.L., Y.S., X.W. continuous-LDSC: A.L., Y.W., Simulations: A.L., Nextflow pipeline: P.A., Interactive Pipeline: Zh.Y., Software support: A.W., X.T., Consultation: J.Y, S.Y, P.F.S, J.H.L, Writing, original draft: A.L., J.Z., N.R.W. Writing, review and editing: A.L., J.Z., N.R.W., P.F.S. Visualization, A.L., J.Z., N.R.W. Supervision: J.Z., N.R.W.

## Data and materials availability

The publicly available data used in this study are available through the references provided in this manuscript or requested through their link. The SC-to-GWAS and GWAS-to-SC benchmarking pipeline is available at https://github.com/Share-AL-work/SC-GWAS_GWAS-SC.

An interactive web portal for visualizing and downloading the benchmark data and results is available at https://gctbhub.cloud.edu.au/shiny/CATCH.

The CATCH pipeline is available at https://github.com/PeterCAllen/CATCH-pipeline.

## Reference

1. Morris, J. A. et al. Discovery of target genes and pathways at GWAS loci by pooled single-cell CRISPR screens. Science (1979). 380, (2023).

2. Yazar, S. et al. Single-cell eQTL mapping identifies cell type-specific genetic control of autoimmune disease. Science 376, (2022).

3. Werder, R. B. et al. CRISPR interference interrogation of COPD GWAS genes reveals the functional significance of desmoplakin in iPSC-derived alveolar epithelial cells. Sci. Adv. 8, (2022).

4. Onuora, S. Single-cell RNA sequencing sheds light on cell-type specific gene expression in immune cells. Nat. Rev. Rheumatol. 18, 363 (2022).

5. Cuomo, A. S. E., Nathan, A., Raychaudhuri, S., MacArthur, D. G. & Powell, J. E. Single-cell genomics meets human genetics. Nature Reviews Genetics 2023 24:8 24, 535–549 (2023).

6. Heath, J. R., Ribas, A. & Mischel, P. S. Single-cell analysis tools for drug discovery and development. Nature Reviews Drug Discovery 2015 15:3 15, 204–216 (2015).

7. Mosquera, J. V. et al. Integrative single-cell meta-analysis reveals disease-relevant vascular cell states and markers in human atherosclerosis. Cell Rep. 42, (2023).

8. Masuda, T., Sankowski, R., Staszewski, O. & Prinz, M. Microglia Heterogeneity in the Single-Cell Era. Cell Rep. 30, 1271–1281 (2020).

9. Zhang, M. J. et al. Polygenic enrichment distinguishes disease associations of individual cells in single-cell RNA-seq data. Nature Genetics 2022 54:10 54, 1572–1580 (2022).

10. Finucane, H. K. et al. Partitioning heritability by functional annotation using genome-wide association summary statistics. Nature Genetics 2015 47:11 47, 1228–1235 (2015).

11. Gazal, S. et al. Linkage disequilibrium–dependent architecture of human complex traits shows action of negative selection. Nature Genetics 2017 49:10 49, 1421–1427 (2017).

12. Skene, N. G. et al. Genetic identification of brain cell types underlying schizophrenia. Nature Genetics 2018 50:6 50, 825–833 (2018).

13. Timshel, P. N., Thompson, J. J. & Pers, T. H. Genetic mapping of etiologic brain cell types for obesity. Elife 9, 1–45 (2020).

14. Jagadeesh, K. A. et al. Identifying disease-critical cell types and cellular processes by integrating single-cell RNA-sequencing and human genetics. Nature Genetics 2022 54:10 54, 1479–1492 (2022).

15. Ma, Y. et al. Polygenic regression uncovers trait-relevant cellular contexts through pathway activation transformation of single-cell RNA sequencing data. Cell Genomics 3, 100383 (2023).

16. Shang, L., Smith, J. A. & Zhou, X. Leveraging gene co-expression patterns to infer trait-relevant tissues in genome-wide association studies. PLoS Genet. 16, e1008734 (2020).

17. Wang, R., Lin, D. Y. & Jiang, Y. EPIC: Inferring relevant cell types for complex traits by integrating genome-wide association studies and single-cell RNA sequencing. PLoS Genet. 18, e1010251 (2022).

18. Calderon, D. et al. Inferring Relevant Cell Types for Complex Traits by Using Single-Cell Gene Expression. Am. J. Hum. Genet. 101, 686–699 (2017).

19. Watanabe, K., Taskesen, E., Van Bochoven, A. & Posthuma, D. Functional mapping and annotation of genetic associations with FUMA. Nature Communications 2017 8:1 8, 1–11 (2017).

20. De Leeuw, C. A., Neale, B. M., Heskes, T. & Posthuma, D. The statistical properties of gene-set analysis. Nat. Rev. Genet. 17, (2016).

21. Visscher, P. M. et al. 10 Years of GWAS Discovery: Biology, Function, and Translation. Am. J. Hum. Genet. 101, 5 (2017).

22. Watanabe, K. et al. A global overview of pleiotropy and genetic architecture in complex traits. Nature Genetics 2019 51:9 51, 1339–1348 (2019).

23. Luecken, M. D. et al. Benchmarking atlas-level data integration in single-cell genomics. Nature Methods 2021 19:1 19, 41–50 (2021).

24. Kim, M. C. et al. Method of moments framework for differential expression analysis of single-cell RNA sequencing data. Cell 187, 6393–6410.e16 (2024).

25. Pullin, J. M. & McCarthy, D. J. A comparison of marker gene selection methods for single-cell RNA sequencing data. Genome Biol. 25, 1–37 (2024).

26. Argelaguet, R., Cuomo, A. S. E., Stegle, O. & Marioni, J. C. Computational principles and challenges in single-cell data integration. Nature Biotechnology 2021 39:10 39, 1202–1215 (2021).

27. Skene, N. G. & Grant, S. G. N. Identification of vulnerable cell types in major brain disorders using single cell transcriptomes and expression weighted cell type enrichment. Front. Neurosci. 10, 179460 (2016).

28. Kim, H. J. et al. Uncovering cell identity through differential stability with Cepo. Nature Computational Science 2021 1:12 1, 784–790 (2021).

29. Dougherty, J. D., Schmidt, E. F., Nakajima, M. & Heintz, N. Analytical approaches to RNA profiling data for the identification of genes enriched in specific cells. Nucleic Acids Res. 38, 4218–4230 (2010).

30. Zeisel, A. et al. Molecular Architecture of the Mouse Nervous System. Cell 174, 999–1014.e22 (2018).

31. de Leeuw, C. A., Mooij, J. M., Heskes, T. & Posthuma, D. MAGMA: Generalized Gene-Set Analysis of GWAS Data. PLoS Comput. Biol. 11, e1004219 (2015).

32. de Leeuw, C. A., Stringer, S., Dekkers, I. A., Heskes, T. & Posthuma, D. Conditional and interaction gene-set analysis reveals novel functional pathways for blood pressure. Nature Communications 2018 9:1 9, 3768-(2018).

33. Xiao, H. et al. STELLA: A Multimodal LLM for Protein Functional Annotation via Unified Sequence-Structure Encoding. Findings of the Association for Computational Linguistics: ACL 2026 25030–25049 (2026) doi:10.18653/V1/2026.FINDINGS-ACL.1254.

34. Nair, S. et al. Agentic systems are adept at solving well-scoped, verifiable problems in computational biology. bioRxiv 2026.04.06.716850 (2026) doi:10.64898/2026.04.06.716850.

35. Chen, S. F. et al. LLM-assisted systematic review of large language models in clinical medicine. Nature Medicine 2026 32:3 32, 1152–1159 (2026).

36. Brodeur, P. G. et al. Superhuman performance of a large language model on the reasoning tasks of a physician. Science (1979). 10.1126/SCIENCE.ADZ4433;PAGEGROUP:STRING:PUBLICATION (2024) doi:10.1126/SCIENCE.ADZ4433;PAGEGROUP:STRING:PUBLICATION.

37. Liu, Y. et al. ACAT: A Fast and Powerful p Value Combination Method for Rare-Variant Analysis in Sequencing Studies. Am. J. Hum. Genet. 104, (2019).

38. Liu, Y. & Xie, J. Cauchy Combination Test: A Powerful Test With Analytic p-Value Calculation Under Arbitrary Dependency Structures. J. Am. Stat. Assoc. 115, 393–402 (2020).

39. Jones, R. C. et al. The Tabula Sapiens: A multiple-organ, single-cell transcriptomic atlas of humans. Science 376, (2022).

40. Fasolino, M. et al. Single-cell multi-omics analysis of human pancreatic islets reveals novel cellular states in type 1 diabetes. Nature Metabolism 2022 4:2 4, 284–299 (2022).

41. Elgamal, R. M. et al. An Integrated Map of Cell Type–Specific Gene Expression in Pancreatic Islets. Diabetes 72, 1719–1728 (2023).

42. Muraro, M. J. et al. A Single-Cell Transcriptome Atlas of the Human Pancreas. Cell Syst. 3, 385–394.e3 (2016).

43. Lai, Q., Dannenfelser, R., Roussarie, J. P. & Yao, V. Disentangling associations between complex traits and cell types with seismic. Nature Communications 2025 16:1 16, 8744-(2025).

44. Finucane, H. K. et al. Heritability enrichment of specifically expressed genes identifies disease-relevant tissues and cell types. Nat. Genet. 50, 621 (2018).

45. Li, A. et al. mBAT-combo: A more powerful test to detect gene-trait associations from GWAS data. The American Journal of Human Genetics 110, 30–43 (2023).

46. Schaum, N. et al. Single-cell transcriptomics of 20 mouse organs creates a Tabula Muris. Nature 2018 562:7727 562, 367–372 (2018).

47. Smillie, C. S. et al. Intra- and Inter-cellular Rewiring of the Human Colon during Ulcerative Colitis. Cell 178, 714–730.e22 (2019).

48. Ward, L. D. & Kellis, M. Interpreting noncoding genetic variation in complex traits and human disease. Nature Biotechnology 2012 30:11 30, 1095–1106 (2012).

49. Mattick, J. S. et al. Long non-coding RNAs: definitions, functions, challenges and recommendations. Nature Reviews Molecular Cell Biology 2023 24:6 24, 430–447 (2023).

50. French, J. D. & Edwards, S. L. The Role of Noncoding Variants in Heritable Disease. Trends in Genetics 36, (2020).

51. Song, L., Chen, W., Hou, J., Guo, M. & Yang, J. Spatially resolved mapping of cells associated with human complex traits. Nature 2025 641:8064 641, 932–941 (2025).

52. Yu, F. et al. Variant to function mapping at single-cell resolution through network propagation. Nature Biotechnology 2022 40:11 40, 1644–1653 (2022).

53. Thrupp, N. et al. Single-Nucleus RNA-Seq Is Not Suitable for Detection of Microglial Activation Genes in Humans. Cell Rep. 32, 108189 (2020).

54. Rouhana, J. M., et al. ECLIPSER: identifying causal cell types and genes for complex traits through single cell enrichment of e/sQTL-mapped genes in GWAS loci. bioRxiv 2021.11.24.469720 (2021) doi:10.1101/2021.11.24.469720.

55. Li, Y. E. et al. A comparative atlas of single-cell chromatin accessibility in the human brain. Science 382, eadf7044 (2023).

56. Farbehi, N. et al. Integrating population genetics, stem cell biology and cellular genomics to study complex human diseases. Nature Genetics 2024 56:5 56, 758–766 (2024).

57. Zhou, Y. et al. scPrediXcan integrates deep learning methods and single-cell data into a cell-type-specific transcriptome-wide association study framework. Cell Genomics 5, 100875 (2025).

58. Zhou, Y. et al. Protocol to perform cell-type-specific transcriptome-wide association study using scPrediXcan framework. STAR Protoc. 7, 104306 (2026).

